# Serum Autotaxin is a Prognostic Indicator of Liver-related Events in Patients with Non-alcoholic Fatty Liver Disease

**DOI:** 10.1101/2023.07.18.23292813

**Authors:** Takanobu Iwadare, Takefumi Kimura, Taiki Okumura, Shun-ichi Wakabayashi, Taro Nakajima, Shohei Kondo, Hiroyuki Kobayashi, Yuki Yamashita, Ayumi Sugiura, Naoyuki Fujimori, Tomoo Yamazaki, Hideo Kunimoto, Satoshi Shimamoto, Koji Igarashi, Satoru Joshita, Naoki Tanaka, Takeji Umemura

## Abstract

**Background:** Circulating autotaxin (ATX) levels have been reported to correlate with liver inflammation activity and liver fibrosis severity in patients with non-alcoholic fatty liver disease (NAFLD). The objective of this study was to investigate whether serum ATX could predict liver-related events (LRE) in NAFLD patients.

**Methods:** This retrospective investigation included 309 biopsy-proven NAFLD patients registered at Shinshu University Hospital between 1998 and 2021. All patients were followed for at least 1 year, during which time the prevalence of LRE, including newly developing hepatocellular carcinoma, hepatic encephalopathy, ascites, and esophagogastric varices, was investigated in relation to ATX levels at the time of liver biopsy.

**Results:** During the median follow-up period of 7.0 years, LRE were observed in 20 patients (6.5%). The area under the receiver operating characteristic curve and cut-off value of serum ATX for predicting LRE were 0.81 and 1.227 mg/L, respectively. Patients with serum ATX greater than the cut-off at the time of liver biopsy had a significantly higher cumulative incidence of LRE than those without (18.4% vs. 2.7%, p < 0.00001). Multivariate Cox proportional hazards models for LRE determined fibrosis stage and ATX divided by the upper limit of normal as independently associated factors.

**Conclusion:** Serum ATX may serve as a predictive marker for LRE in patients with NAFLD.

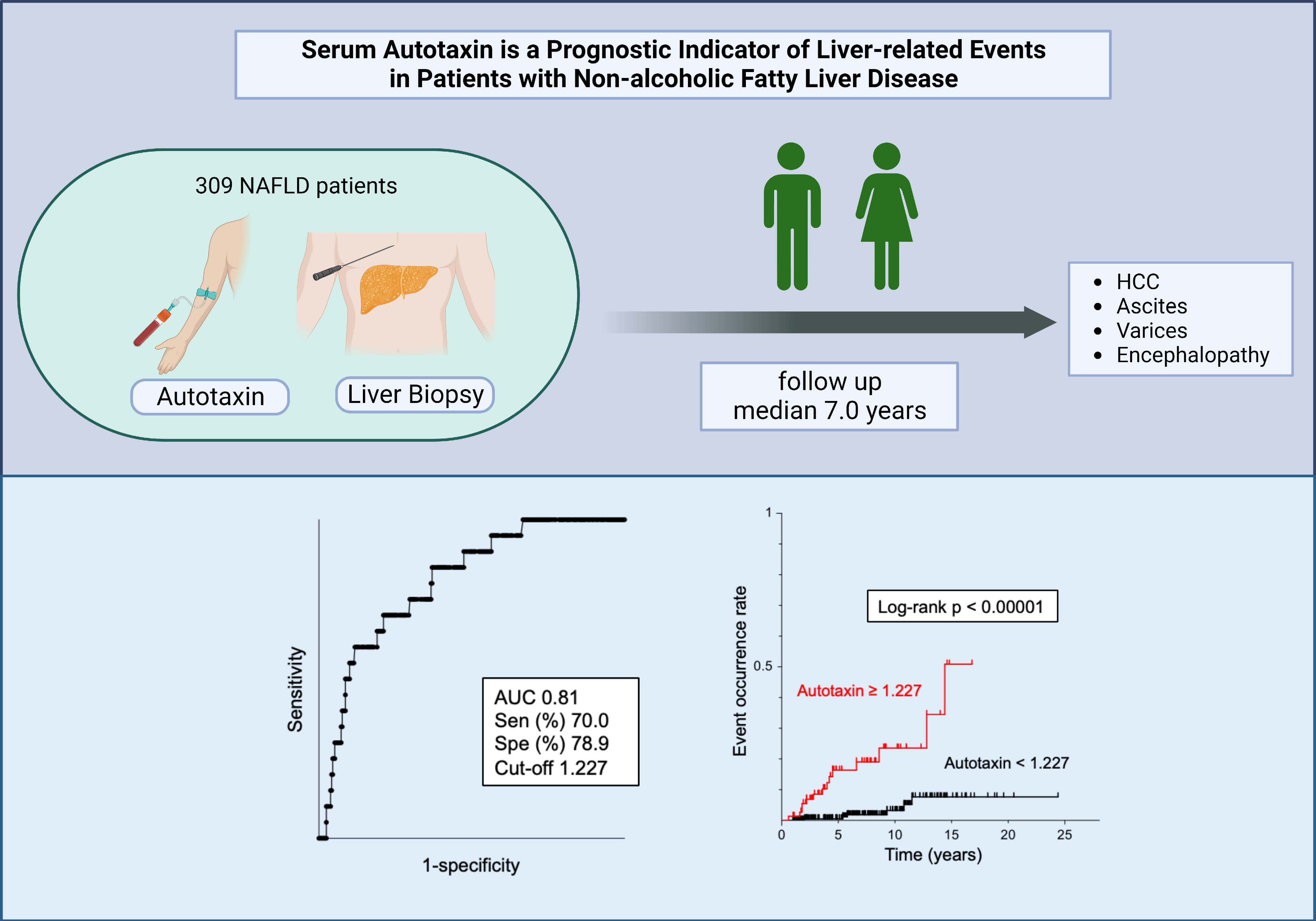

## Introduction

As a hepatic manifestation of metabolic syndrome, non-alcoholic fatty liver disease (NAFLD) is closely linked to obesity, hypertension (HT), diabetes mellitus (DM), and dyslipidemia (DL),1 and is increasing worldwide.2 NAFLD is categorized as non-alcoholic fatty liver and non-alcoholic steatohepatitis, the latter of which raises the risk of cirrhosis, liver failure, and hepatocellular carcinoma (HCC).3 In patients with NAFLD, liver fibrosis is considered the best indicator of long-term clinical prognosis,4 5 and so the establishment of accurate, minimally invasive, and safe markers of liver fibrosis are desired.6 7 8

Autotaxin (ATX) was originally discovered in human melanoma cell cultures.9 The protein is encoded by the ectonucleotide pyrophosphatase/phosphodiesterase family member 2 gene, which catalyzes the hydrolysis of lysophosphatidylcholine to lysophosphatidic acid (LPA) and functions as a phospholipase.10 We have shown that serum ATX levels correlate with liver inflammation activity and fibrosis severity in viral hepatitis, primary biliary cholangitis, and NAFLD.11 12 13 14 Since 2018, serum ATX measurement has been covered by Japanese national health insurance for patients with chronic hepatitis and liver cirrhosis, and its clinical application is already established. However, no studies have evaluated the significance of ATX as a predictor of NAFLD outcome. We therefore investigated whether circulating ATX levels reflected the risk of liver-related events (LRE) in NAFLD patients.

## Methods

### Patients and clinical examinations

This retrospective study was approved by the Committee for Medical Ethics of Shinshu University School of Medicine (ID number: 4285) and performed in accordance with the Helsinki declaration of 1975, 1983 revision. We firstly enrolled 409 biopsy-proven Japanese NAFLD patients who were admitted to Shinshu University Hospital (Matsumoto, Japan) between January 1998 and September 2021. NAFLD was suspected based on the following criteria: 1 the presence of hepatorenal contrast and increased hepatic echogenicity on abdominal ultrasonography; 2 an average daily consumption of < 20 g of ethanol; and 3 the absence of other causes of liver dysfunction, such as viral hepatitis, drug-induced liver injury, autoimmune liver disease, primary sclerosing cholangitis, Wilson’s disease, hereditary hemochromatosis, and citrin deficiency.15 The diagnosis of NAFLD was confirmed using the histological findings of biopsied specimens. Of the liver biopsy-diagnosed 409 NAFLD patients, 384 had samples available at the time of liver biopsy, among which 309 were followed for at least 1 year and included in the study (Figure 1).

**Figure 1.**
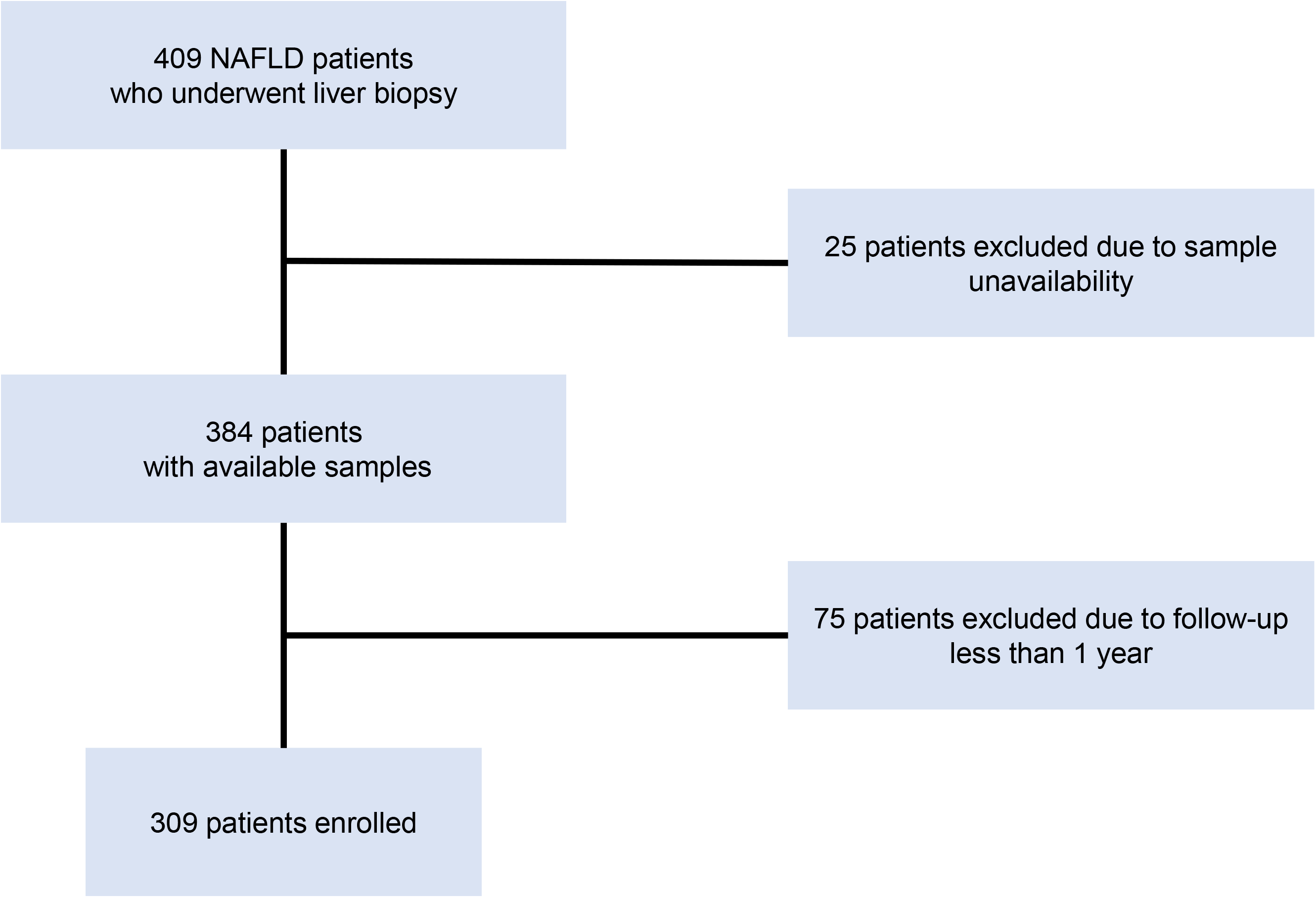
Patient inclusion flowchart for this study. NAFLD, non-alcoholic fatty liver disease

Patients were considered to have HT if their systolic/diastolic pressure was > 140/90 mmHg or if they were taking anti-hypertensive drugs.16 Patients were judged as having HL if their fasting serum levels of total cholesterol (TC), low density lipoprotein cholesterol (LDL-C) or triglycerides were ≥ 220 mg/dL, ≥ 140 mg/dL, or ≥ 150 mg/dL, respectively, or if they were taking lipid-lowering drugs.17 Patients were considered to have DM with a fasting glucose level of ≥ 126 mg/dL or hemoglobin A1c of ≥ 6.5%, or if they were taking insulin or oral hypoglycemic agents.18 Body weight and height were measured before liver biopsy in an overnight fasting state. All laboratory data were obtained in an overnight fasting state on the day of liver biopsy. Serum levels of ATX were determined using a two-site enzyme immunoassay and an automated immunoassay analyzer (Tosoh Corporation, Tokyo, Japan). Homeostasis model assessment for insulin resistance (HOMA-IR) was calculated according to the following formula: HOMA-IR = (fasting blood glucose [mg/dL] x fasting insulin [μU/mL])/405.19

### Histological examinations

Liver specimens of at least 1.5 cm in length were obtained from segments 5 or 8 using a 14-gauge needle as described previously and immediately fixed in 10% neutral formalin.8 Sections of 4 μm in thickness were cut and stained using the hematoxylin and eosin and Azan-Mallory methods. The histological activity of NAFLD was assessed by an independent expert pathologist in a blinded manner according to the established NAFLD scoring system.20 Steatosis was graded as 0 to 3 based on the rate of steatotic hepatocytes (< 5%, 5-33%, > 33-66%, and > 66%, respectively). Lobular inflammation was graded as 0 to 3 based on the overall assessment of all inflammatory foci (no foci, < 2 foci/200× field, 2-4 foci/200× field, and > 4 foci/200× field, respectively). Ballooning grade was scored as 0-2 by the frequency of ballooned hepatocytes (none, few, and many, respectively). Fibrosis stage was scored as follows: F0, none; F1, perisinusoidal or periportal; F2, perisinusoidal and portal/periportal; F3, bridging fibrosis; and F4, cirrhosis.

### Patient follow-up

The 309 analyzed patients were monitored every 6 months by ultrasonography or computed tomography and measurement of serum alpha-fetoprotein (AFP), with cirrhotic patients evaluated every 3 months. The radiological diagnosis of HCC was based on the American Association for the Study of Liver Diseases practice guidelines on the management of HCC as either: 1 the presence of a hepatic lesion > 2 cm in diameter with typical vascular pattern for HCC on one dynamic imaging technique or AFP > 200 ng/mL; or 2 the presence of a lesion 1-2 cm in diameter with typical vascular pattern for HCC on two dynamic imaging techniques.21 LRE were defined as the development of HCC, hepatic encephalopathy of grade II or higher, poorly controlled ascites requiring hospitalization, and esophagogastric varices requiring endoscopic ligation, sclerotherapy, and balloon-occluded retrograde transvenous obliteration, including varices rupture. Follow-up time was defined as the number of years from biopsy to event diagnosis or from biopsy to the last follow-up visit when protocol surveillance confirmed no event.

### Statistical analysis

Clinical data were expressed as the number (percentage) or as the median (interquartile range [IQR]). Statistical analyses were performed using StatFlex Ver. 7.0 and R software ver. 4.3.0. The Mann–Whitney U test and Chi-square test were employed for comparisons between the study groups. Diagnostic accuracy was evaluated using the area under the receiver operating characteristic curve (AUROC). The Youden index identified cut-off values, with the nearest clinically applicable value to the cut-off considered the optimal threshold for clinical convenience. The Kaplan–Meier method and log-rank testing were used to estimate disease progression. The Cox proportional hazards model was adopted to assess univariate and multivariable covariates for LRE. All statistical tests were two-tailed and evaluated at the 0.05 level of significance.

## Results

### Baseline characteristics

The clinicopathological features of the 309 patients with NAFLD who were monitored for more than 1 year are presented in Table 1. Median age at the time of biopsy was 56 years, and 135 patients (44%) were male. The elevated complication rates of DM (39%), HT (42%), and DL (63%) were typical for a NAFLD population. The median values for body mass index, aspartate aminotransferase, alanine aminotransferase, and HOMA-IR were 26.6 kg/m^2^, 49 U/L, 71 U/L, and 3.5, respectively. Median serum ATX was 0.87 mg/L. The histopathological classification by steatosis grade 1/2/3 was 96/138/75 patients, respectively. Similarly, respective lobular inflammation grade 0/1/2/3 was 14/151/127/17 patients, and ballooning grade 0/1/2 was 53/168/88 patients. According to fibrosis stage F0, F1, F2, F3, and F4, the number of patients in each stage was 46, 136, 37, 70, and 20, respectively.

**Table 1.**
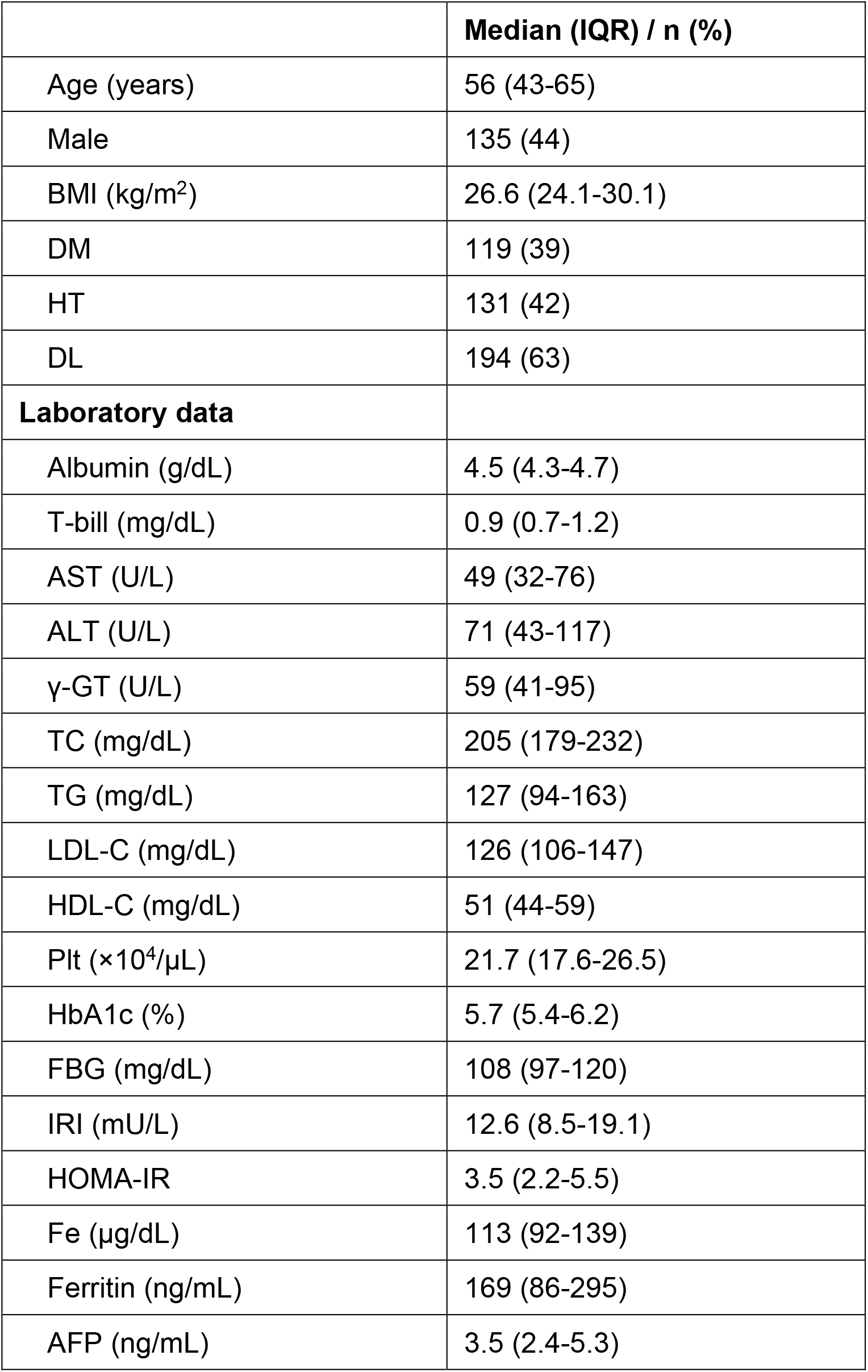
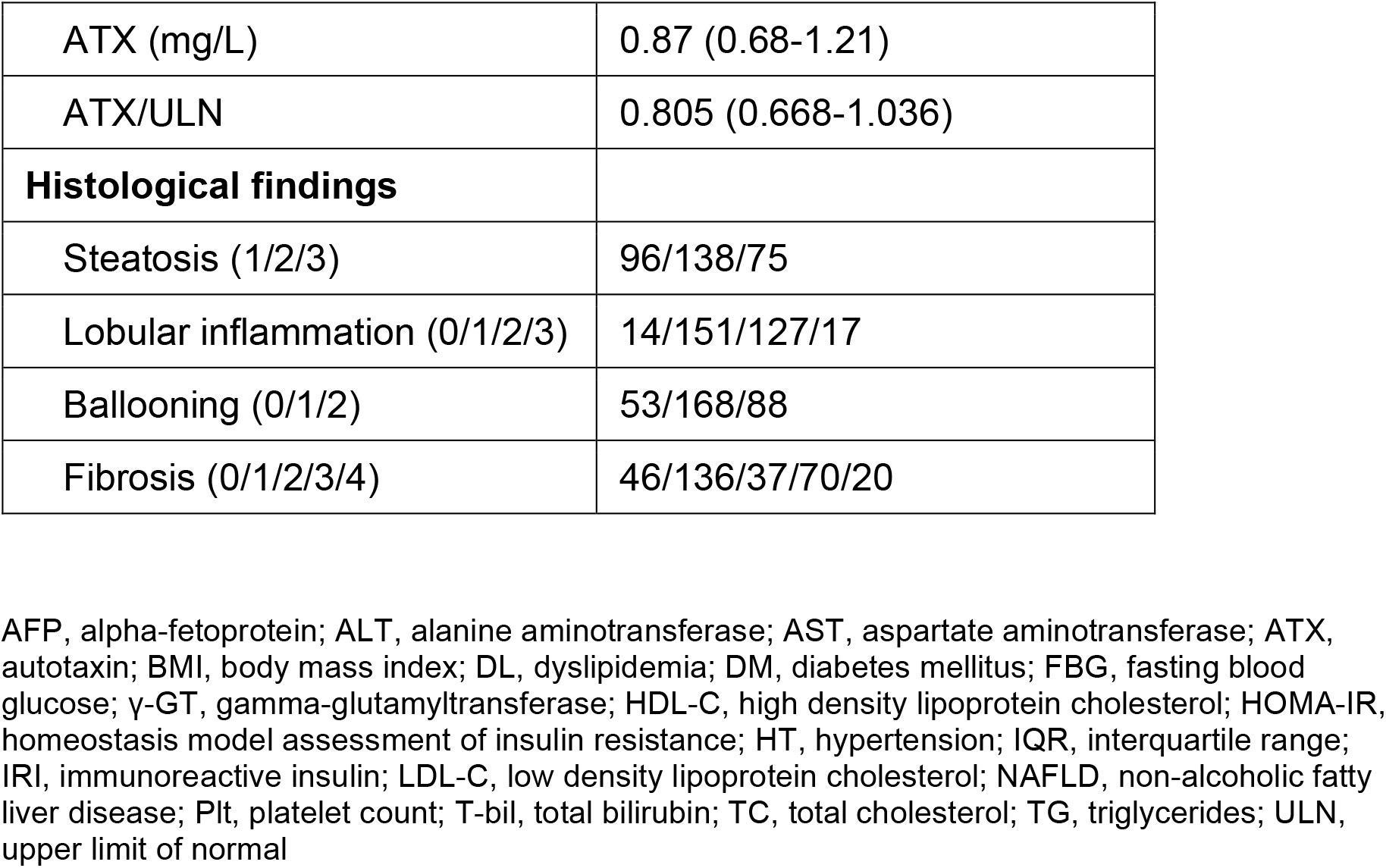
Baseline characteristics of 309 patients with NAFLD.

### Occurrence of events

The median follow-up evaluation period for the 309 patients was 7.0 years (IQR: 3.8-10.1 years). A total of 12 patients (3.9%; 4 male and 8 female) reached the outcome of death. Liver-related death occurred in 7 patients: liver failure in 4 patients, HCC in 2 patients, and ruptured varices in 1 patient. The cause of death in the remaining 5 patients were diverse (breast cancer, heart failure, renal failure, acute myeloid leukemia, and unknown in 1 patient each). Nine (2.9%) and 20 (6.5%) patients newly developed HCC and LRE, respectively (Table 2).

**Table 2.** Details of event occurrences.

|  | **All (n = 309)** | **Male (n = 135)** | **Female (n = 174)** |
| --- | --- | --- | --- |
|  | **Median (IQR) / n (%)** | **Median (IQR) / n (%)** | **Median (IQR) / n (%)** |
| Follow-up (years) | 7.0 (3.8-10.1) | 6.7 (3.4-9.5) | 7.1 (3.8-10.2) |
| Events during follow-up |  |  |  |
| Death | 12 (3.9) | 4 (3.0) | 8 (4.6) |
| Hepatocellular carcinoma | 9 (2.9) | 4 (2.9) | 5 (2.9) |
| Liver-related event | 20 (6.5) | 7 (5.1) | 13 (7.5) |

IQR, interquartile range

### Comparison of clinicopathologic features between non-LRE and LRE patients

To identify the predictors of LRE, clinicopathological features at the time of biopsy were compared between non-LRE and LRE patients (Table 3). Patients with LRE had significantly higher age (p < 0.001), higher prevalences of DM (p = 0.012) and HT (p < 0.001), higher levels of fasting blood glucose (p = 0.046), insulin (p = 0.049), and ATX (p < 0.001), lower prevalence of DL (p = 0.009), and lower levels of albumin (p < 0.001), TC (p < 0.001), LDL-C (p < 0.001), and platelet count (p < 0.001) compared with non-LRE patients. Regarding pathological findings, LRE patients had a lower steatosis score (p = 0.003) along with a higher lobular inflammation score (p = 0.013) and fibrosis stage (p < 0.001) than did non-LRE patients.

**Table 3.**
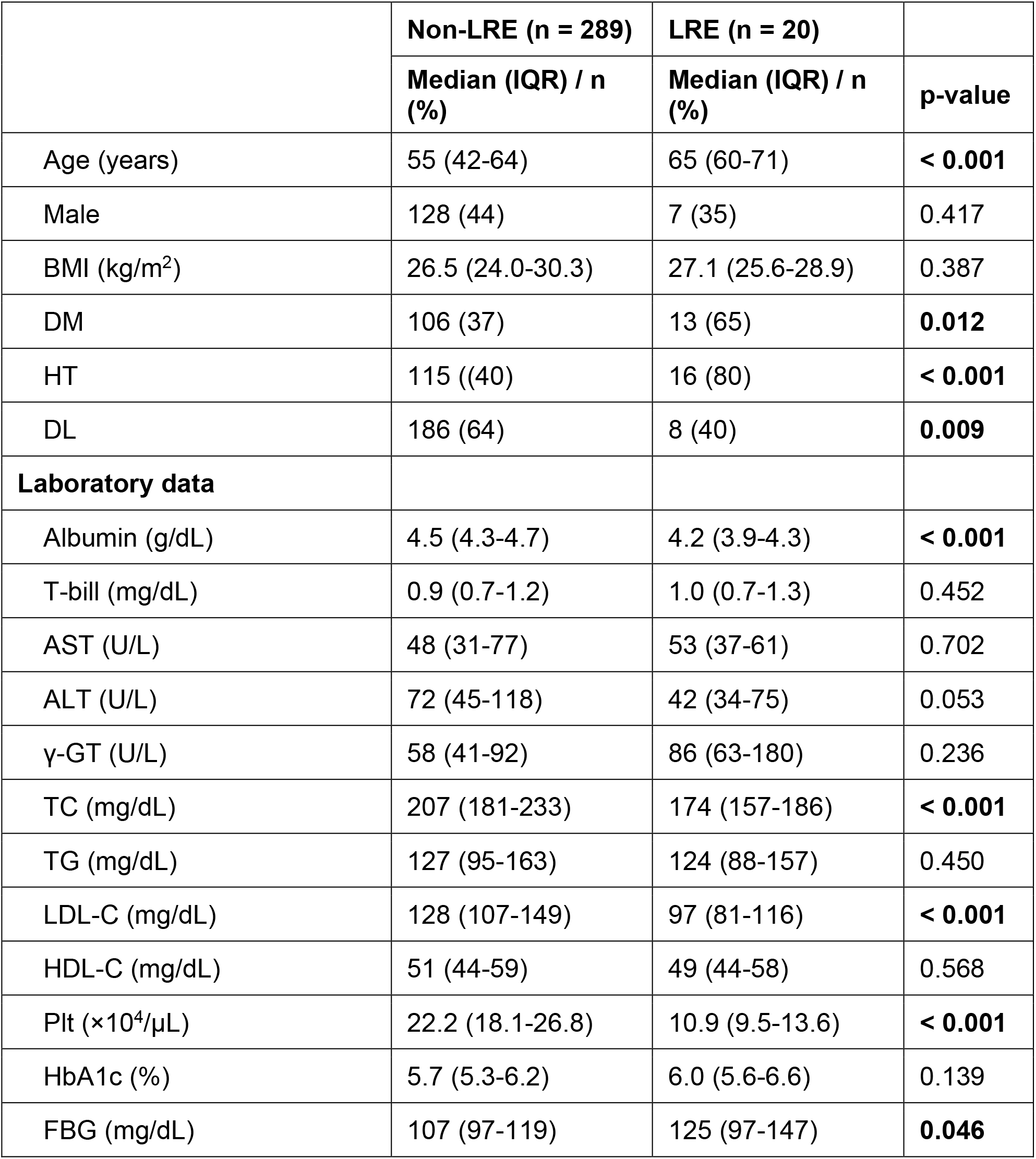
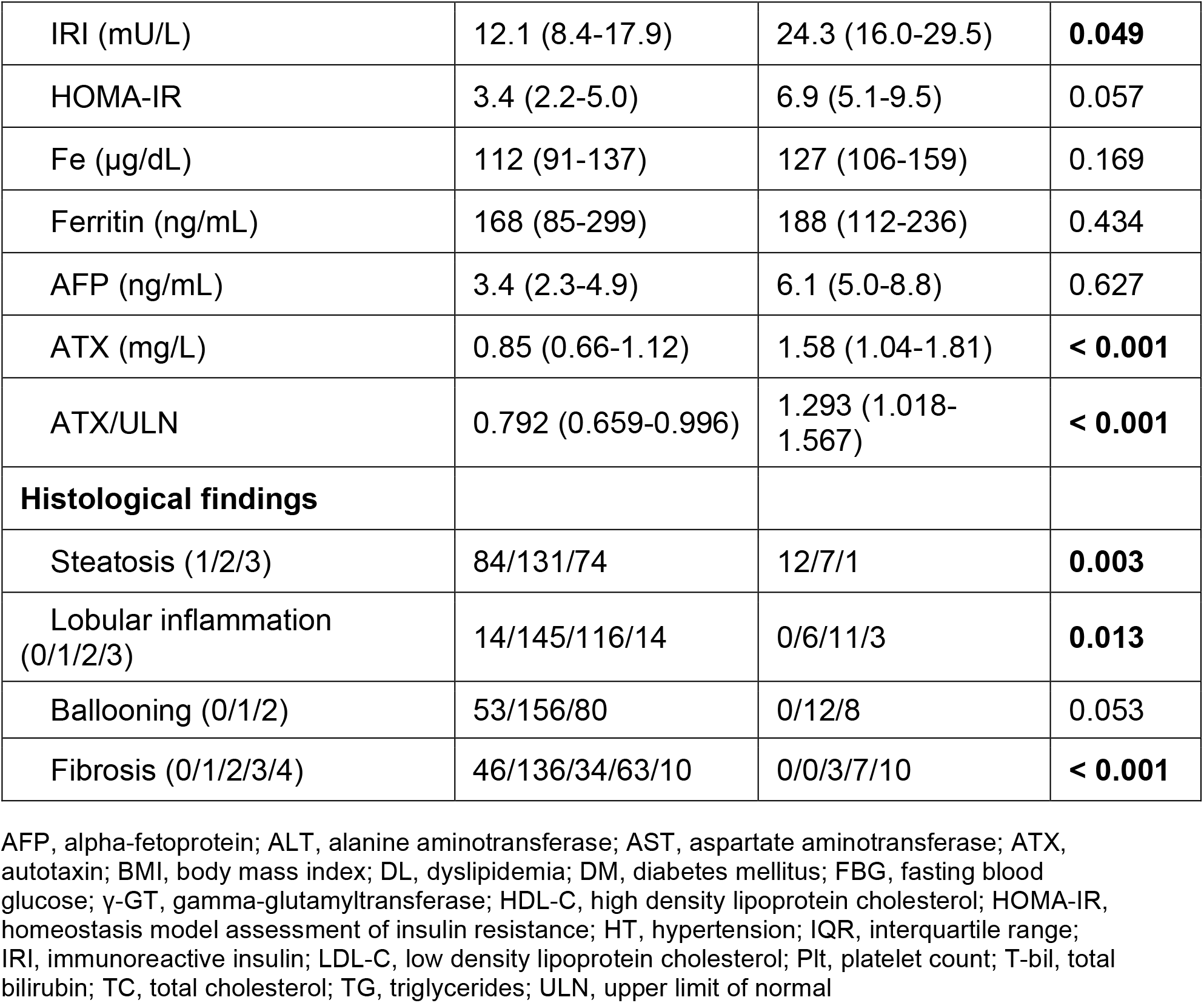
Comparisons of clinicopathological features at time of biopsy between non-LRE and LRE patients.

|  | **Non-LRE (n = 289)** | **LRE (n = 20)** |  |
| --- | --- | --- | --- |
|  | **Median (IQR) / n (%)** | **Median (IQR) / n (%)** | **p-value** |
| Age (years) | 55 (42-64) | 65 (60-71) | **< 0.001** |
| Male | 128 (44) | 7 (35) | 0.417 |
| BMI (kg/m<sup>2</sup>) | 26.5 (24.0-30.3) | 27.1 (25.6-28.9) | 0.387 |
| DM | 106 (37) | 13 (65) | **0.012** |
| HT | 115 ((40) | 16 (80) | **< 0.001** |
| DL | 186 (64) | 8 (40) | **0.009** |
| **Laboratory data** |  |  |  |
| Albumin (g/dL) | 4.5 (4.3-4.7) | 4.2 (3.9-4.3) | **< 0.001** |
| T-bill (mg/dL) | 0.9 (0.7-1.2) | 1.0 (0.7-1.3) | 0.452 |
| AST (U/L) | 48 (31-77) | 53 (37-61) | 0.702 |
| ALT (U/L) | 72 (45-118) | 42 (34-75) | 0.053 |
| γ-GT (U/L) | 58 (41-92) | 86 (63-180) | 0.236 |
| TC (mg/dL) | 207 (181-233) | 174 (157-186) | **< 0.001** |
| TG (mg/dL) | 127 (95-163) | 124 (88-157) | 0.450 |
| LDL-C (mg/dL) | 128 (107-149) | 97 (81-116) | **< 0.001** |
| HDL-C (mg/dL) | 51 (44-59) | 49 (44-58) | 0.568 |
| Plt (×10<sup>4</sup>/μL) | 22.2 (18.1-26.8) | 10.9 (9.5-13.6) | **< 0.001** |
| HbA1c (%) | 5.7 (5.3-6.2) | 6.0 (5.6-6.6) | 0.139 |
| FBG (mg/dL) | 107 (97-119) | 125 (97-147) | **0.046** |
| IRI (mU/L) | 12.1 (8.4-17.9) | 24.3 (16.0-29.5) | **0.049** |
| HOMA-IR | 3.4 (2.2-5.0) | 6.9 (5.1-9.5) | 0.057 |
| Fe (µg/dL) | 112 (91-137) | 127 (106-159) | 0.169 |
| Ferritin (ng/mL) | 168 (85-299) | 188 (112-236) | 0.434 |
| AFP (ng/mL) | 3.4 (2.3-4.9) | 6.1 (5.0-8.8) | 0.627 |
| ATX (mg/L) | 0.85 (0.66-1.12) | 1.58 (1.04-1.81) | **< 0.001** |
| ATX/ULN | 0.792 (0.659-0.996) | 1.293 (1.018-1.567) | **< 0.001** |
| **Histological findings** |  |  |  |
| Steatosis (1/2/3) | 84/131/74 | 12/7/1 | **0.003** |
| Lobular inflammation (0/1/2/3) | 14/145/116/14 | 0/6/11/3 | **0.013** |
| Ballooning (0/1/2) | 53/156/80 | 0/12/8 | 0.053 |
| Fibrosis (0/1/2/3/4) | 46/136/34/63/10 | 0/0/3/7/10 | **< 0.001** |

AFP, alpha-fetoprotein; ALT, alanine aminotransferase; AST, aspartate aminotransferase; ATX, autotaxin; BMI, body mass index; DL, dyslipidemia; DM, diabetes mellitus; FBG, fasting blood glucose; γ-GT, gamma-glutamyltransferase; HDL-C, high density lipoprotein cholesterol; HOMA-IR, homeostasis model assessment of insulin resistance; HT, hypertension; IQR, interquartile range; IRI, immunoreactive insulin; LDL-C, low density lipoprotein cholesterol; Plt, platelet count; T-bil, total bilirubin; TC, total cholesterol; TG, triglycerides; ULN, upper limit of normal

### Cumulative event incidence rate for entire cohort

Based on the receiver operating characteristic analysis, we determined serum ATX cut-off values for the outcomes of mortality, HCC, and LRE (Figure 2A-C). The cut-off value for predicting death was 1.227 mg/L, with corresponding values for AUROC, sensitivity, and specificity of 0.78, 75.0%, and 77.8%, respectively. Similarly for the prediction of HCC occurrence, the cut-off value was identified as 1.055 mg/L, yielding an AUROC of 0.76, sensitivity of 77.8%, and specificity of 68.7%. The cut-off value for LRE occurrence was 1.227 mg/L, with corresponding values for AUROC, sensitivity, and specificity of 0.81, 70.0%, and 78.9%, respectively.

**Figure 2.**
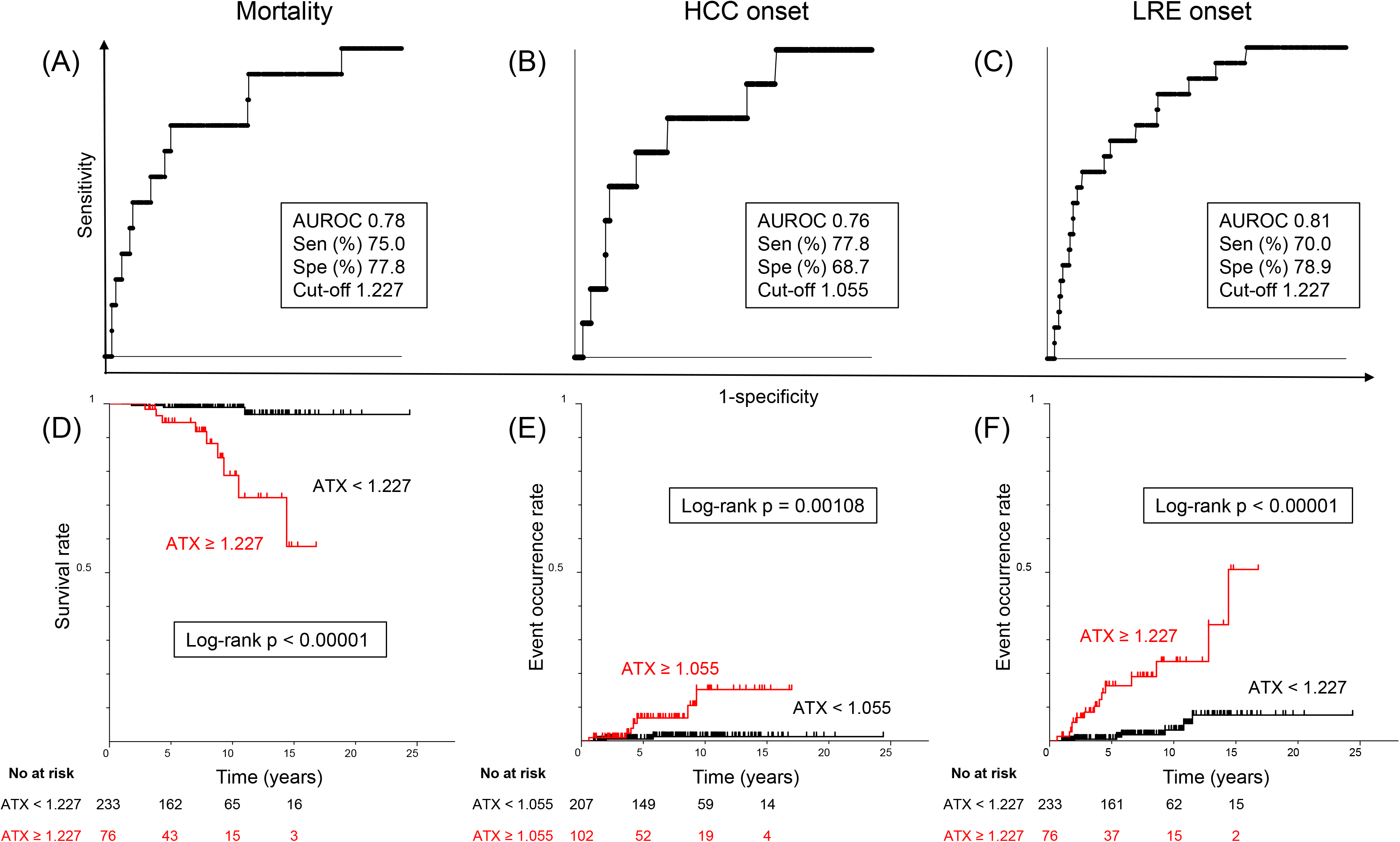
Cumulative event incidence rate analysis of serum ATX levels for entire cohort. (A)(B)(C) Receiver operating characteristic analysis of serum ATX levels for mortality (A), HCC (B), and LRE (C). (D)(E)(F) Cumulative event incidence rate analysis by the Kaplan–Meier method of serum ATX levels for mortality (D), HCC (E), and LRE (F). ATX, autotaxin; AUROC, area under the receiver operating characteristic curve; HCC, hepatocellular carcinoma; LRE, liver-related events; Sen, sensitivity; Spe, specificity

Chi-square testing was performed to compare incidence rates of low-ATX and high-ATX groups as defined by respective cut-off values. The incidence rates for death, HCC, and LRE were all higher in the high-ATX group (11.8% vs. 1.3%; p = 0.00014, 6.9% vs. 1.0%; p = 0.00323, and 18.4% vs. 2.7%; p < 0.00001, respectively).

Kaplan–Meier survival analysis using the cut-off value of 1.227 mg/L revealed a significantly lower survival rate in high-ATX patients than in low-ATX patients (log-rank p < 0.00001). Using the respective cut-off values of 1.055 mg/L and 1.227 mg/L, the cumulative HCC and LRE incidence rates were both higher in high-ATX patients (log rank p = 0.00108 and p < 0.00001, respectively) (Figure 2D-F). The above results indicated that serum ATX levels at the time of biopsy could be a useful parameter for predicting future death and the occurrences of HCC and LRE.

### Cumulative event incidence rate by gender

Previous research has consistently highlighted sex differences in serum ATX levels.22 In agreement with this, we assessed the utility of serum ATX by gender. The optimal cut-off value of ATX for predicting death was 0.875 mg/L for men (AUROC: 0.77, sensitivity: 75.0%, specificity: 79.4%) and 1.214 mg/L for women (AUROC: 0.80, sensitivity: 87.5%, specificity: 64.4%) (Figure 3A, 3D). Kaplan–Meier survival analysis using the respective cut-off values for each gender indicated a significantly lower survival rate among the high-ATX subgroups for both sexes (male: log-rank p = 0.00384, female: log-rank p = 0.00108) (Figure 4A, 4D).

**Figure 3.**
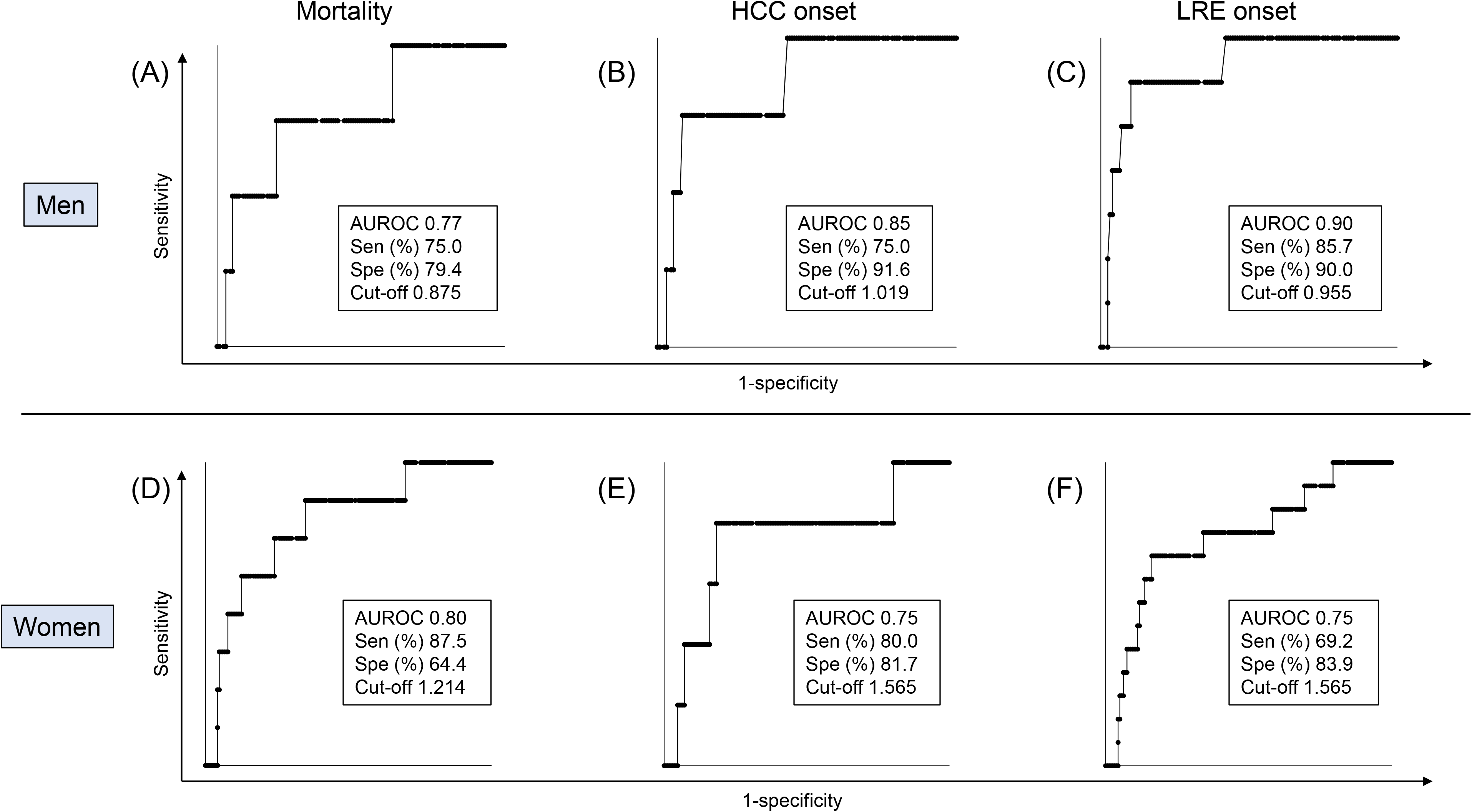
Receiver operating characteristic analysis of serum ATX levels for event occurrence by gender. (A)(B)(C) Receiver operating characteristic analysis of ATX levels for mortality (A), HCC (B), and LRE (C) in men. (D)(E)(F) Receiver operating characteristic analysis of ATX values for mortality (D), HCC (E), and LRE (F) in women. ATX, autotaxin; AUROC, area under the receiver operating characteristic curve; HCC, hepatocellular carcinoma; LRE, liver-related events; Sen, sensitivity; Spe, specificity

**Figure 4.**
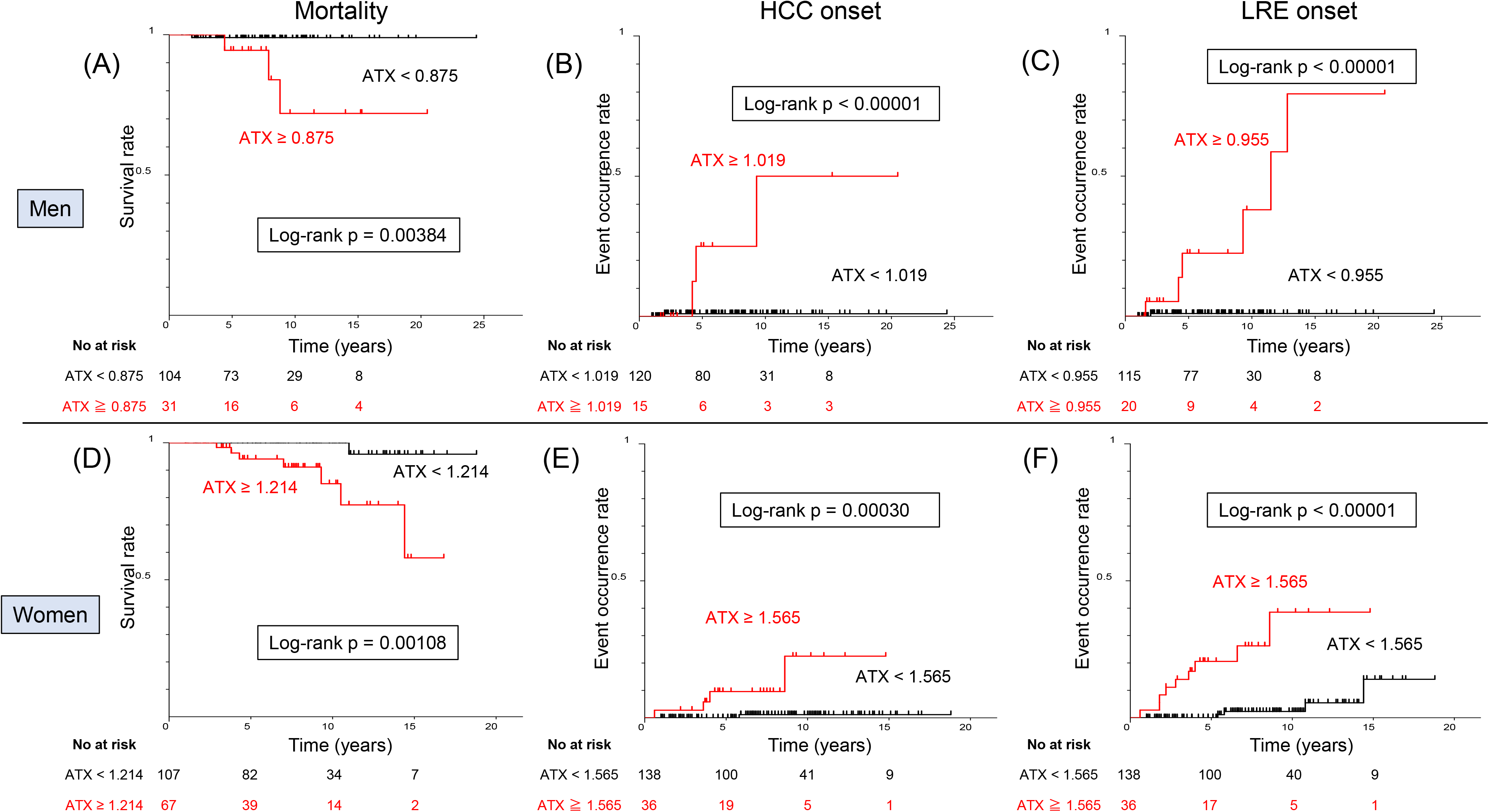
Cumulative event incidence rate analysis of serum ATX levels by gender. (A)(B)(C) Cumulative event incidence rate analysis by the Kaplan–Meier method of serum ATX levels in men for mortality (A), HCC (B), and LRE (C). (D)(E)(F) Cumulative event incidence rate analysis by the Kaplan–Meier method of serum ATX levels in women for mortality (D), HCC (E), and LRE (F). ATX, autotaxin; AUROC, area under the receiver operating characteristic curve; HCC, hepatocellular carcinoma; LRE, liver-related events

Similarly for the onset of HCC, the optimal cut-off value for serum ATX level was 1.019 mg/L for men (AUROC: 0.85, sensitivity: 75.0%, specificity: 91.6%) and 1.565 mg/L for women (AUROC: 0.75, sensitivity: 80.0%, specificity: 81.7%) (Figure 3B, 3E). Kaplan–Meier survival analysis using the respective cut-off values for men and women revealed a significantly higher HCC onset rate in the high-ATX subgroups for both genders (male: log-rank p < 0.00001, female: log-rank p = 0.00030) (Figure 4B, 4E). The optimal cut-off value for serum ATX predicting the onset of LRE was 0.955 mg/L for men (AUROC: 0.90, sensitivity: 85.7%, specificity: 90.0%) and 1.565 mg/L for women (AUROC: 0.75, sensitivity: 69.2%, specificity: 83.9%) (Figure 3C, 3F). Kaplan–Meier survival analysis using the respective cut-off values for men and women showed a significantly higher rate of LRE onset in the high-ATX subgroups for both sexes (male: log-rank p < 0.00001, female: log-rank p < 0.00001) (Figure 4C, 4F). The above results demonstrated the importance of setting cut-off values by gender when using ATX as a predictive marker.

### Cumulative event incidence rate for entire cohort by ATX/upper limit of normal

Using receiver operating characteristic analysis, we calculated cut-off values for serum ATX levels divided by the upper limit of normal (ULN) for ATX in each gender for the outcomes of death, HCC, and LRE (Figure 5A-C). The cut-off value for predicting death was determined as 0.966, with corresponding values for AUROC, sensitivity, and specificity of 0.79, 83.3%, and 71.4%, respectively. For the prediction of HCC, the cut-off value was 1.157, with an AUROC of 0.79, sensitivity of 77.8%, and specificity of 83.7%. The cut-off value for LRE occurrence was 1.063, with corresponding values for AUROC, sensitivity, and specificity of 0.81, 75.0%, and 81.3%, respectively.

**Figure 5.**
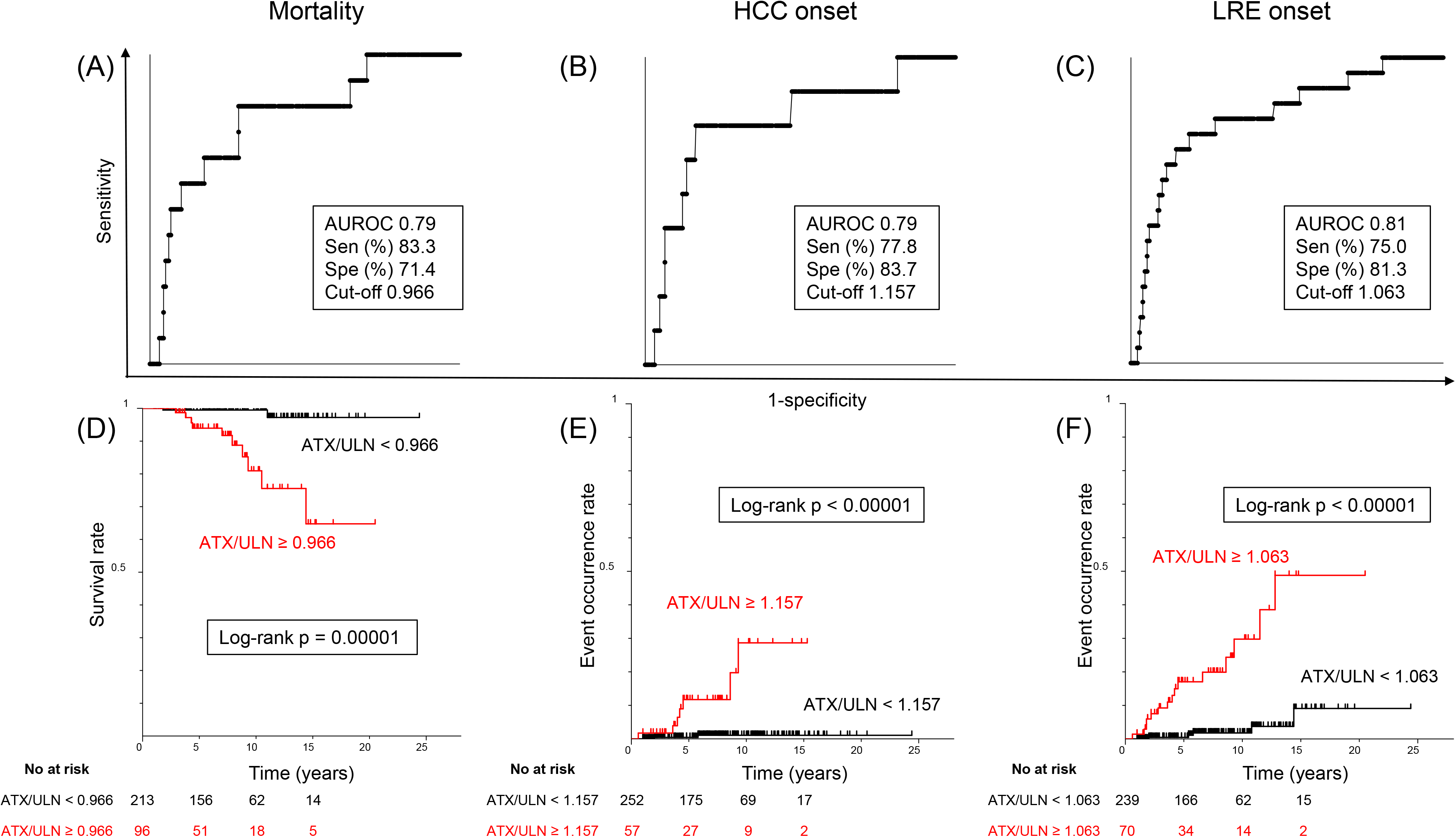
Cumulative event incidence rate analysis for entire cohort of ATX/ULN by gender. (A)(B)(C) Receiver operating characteristic analysis of ATX/ULN for each gender for mortality (A), HCC (B), and LRE (C). (D)(E)(F) Cumulative event incidence rate analysis by the Kaplan–Meier method of ATX/ULN for each gender for mortality (D), HCC (E), and LRE (F). ATX, autotaxin; AUROC, area under the receiver operating characteristic curve; HCC, hepatocellular carcinoma; LRE, liver-related events; Sen, sensitivity; Spe, specificity; ULN, upper limit of normal

Kaplan–Meier survival analysis using the cut-off value for mortality showed that survival rate was significantly lower in high-ATX patients than in low-ATX patients (log-rank p = 0.00001). Using the respective cut-off values of 1.157 and 1.063, the HCC and LRE incidence rates were both significantly higher in high-ATX patients as well (log rank p < 0.00001 and p < 0.00001, respectively) (Figure 5D-F). These results indicated that ATX could be a more useful parameter when evaluated separately for men and women.

### Multivariate Cox proportional hazards models for LRE risk determination

Table 4 summarizes the results of Cox proportional hazards regression analysis for LRE. In univariate Cox proportional hazards testing, age (hazard ratio [HR] 1.089, 95% confidence interval [CI] 1.040-1.141; p < 0.001), HT (HR 5.926, 95% CI 1.976-17.769; p < 0.001), DM (HR 2.633, 95% CI 1.047-6.624; p = 0.040), fibrosis stage (HR 5.239, 95% CI 2.947-9.313; p < 0.001), and ATX/ULN (HR 4.036, 95% CI 2.315-5.750; p < 0.001) were identified as possible determinants of LRE. Three multivariate models were generated using the variables included in the univariate analysis. In all models, ATX/ULN was found to be an independent determinant of LRE occurrence along with fibrosis stage, suggesting that both fibrosis stage and serum ATX expressed as ATX/ULN might serve as important predictive factors for the occurrence of LRE in patients with NAFLD.

**Table 4.** Univariate and multivariate Cox proportional hazards analysis for LRE.

|  | Univariate |  |  | Multivariate Model 1 |  |  | Multivariate Model 2 |  |  | Multivariate Model 3 |  |  |
| --- | --- | --- | --- | --- | --- | --- | --- | --- | --- | --- | --- | --- |
|  | HR | 95% CI of HR | p value | HR | 95% CI of HR | p value | HR | 95% CI of HR | p value | HR | 95% CI of HR | p value |
| Age | 1.089 | 1.040-1.141 | **< 0.001** | 1.036 | 0.977-1.099 | 0.241 | 1.016 | 0.96-1.075 | 0.586 | – | – | – |
| Male | 0.717 | 0.286-1.799 | 0.479 | 2.572 | 0.863-7.669 | 0.09 | – | – | – | 1.947 | 0.707-5.364 | 0.198 |
| HT | 5.926 | 1.976-17.769 | **0.001** | 1.677 | 0.564-4.988 | 0.353 | 2.942 | 0.735-11.781 | 0.127 | 3.94 | 1.115-13.916 | **0.033** |
| DM | 2.633 | 1.047-6.624 | **0.04** | 2.744 | 0.702-10.719 | 0.147 | 1.376 | 0.478-3.965 | 0.554 | 1.692 | 0.567-5.046 | 0.345 |
| Fibrosis stage | 5.239 | 2.947-9.313 | **< 0.001** | 4.331 | 2.23-8.412 | **< 0.001** | 4.373 | 2.252-8.492 | **< 0.001** | 4.715 | 2.486-8.943 | **< 0.001** |
| ATX/ULN | 4.036 | 2.315-5.75 | **< 0.001** | 4.116 | 1.09-15.549 | **0.037** | 3.627 | 1.062-12.384 | **0.04** | 3.927 | 1.132-13.625 | **0.031** |

ATX, autotaxin; CI, confidence interval; DM, diabetes mellitus; HR, hazard ratio; HT, hypertension; ULN, upper limit of normal

## Discussion

The present study evaluated serum ATX in patients with NAFLD to determine its potential to estimate disease prognosis, including mortality and the development of HCC and LRE. Receiver operating characteristic analysis revealed ATX cut-off values of 1.227 mg/L, 1.055 mg/L, and 1.227 mg/L for predicting death, HCC, and LRE, respectively, with corresponding AUROC values of 0.78, 0.76, and 0.81. Log-rank testing showed high cumulative mortality rate (p < 0.00001), HCC incidence rate (p = 0.00108), and LRE incidence rate (p < 0.00001) for each calculated cut-off value, as well as high rates according to gender. Thus, serum ATX exhibited potential utility in predicting such prognostic factors as death, HCC, and LRE in patients with NAFLD. To our knowledge, this is the first report evaluating serum ATX and prognosis in NAFLD.

ATX has been identified as a novel contributor to the pathogenesis of liver fibrosis.23 According to Nakagawa et al., LPA stimulates the proliferation and contractility of hepatic stellate cells, which are the main producers of extracellular matrix components in the liver, with ATX and LPA directly involved in liver fibrosis pathology.22 Another indirect theory exists regarding the relationship between serum ATX and liver fibrosis. In the healthy liver, sinusoidal endothelial cells clear ATX taken up from hepatic sinusoids. However, in fibrotic liver tissue, the capillarization of sinusoids leads to reduced ATX uptake and increased plasma levels.24 Thus, any liver condition that causes fibrosis may result in elevated plasma ATX.

Recent studies have identified a significant correlation between serum ATX levels and fibrosis in NAFLD.11 25 It is well known and confirmed in the present investigation that liver fibrosis is also a relevant factor in the development of LRE.26 However, our finding that ATX/ULN values may predict LRE independently of fibrosis stage is remarkable. Further studies are needed to clarify the detailed mechanism of the relationship between ATX and LRE development. Later in this section, we will consider several hypotheses on the link between ATX and LRE.

ATX is a novel player in the pathogenesis of HCC.23 Along with vascular endothelial growth factor receptor-2 and -3, ATX is reportedly involved in vascular development in the liver, which may contribute to HCC onset during the progression of chronic viral hepatitis C.27 Supporting these results, a recent report indicated that serum ATX levels after antiviral therapy for hepatitis C could predict the formation of HCC during long-term follow-up.28 ATX binds to LPA and adhesion molecules, including integrins, potentially contributing to the metastasis of cancer cells.29 In addition to a role in HCC development, ATX and LPA signaling have been implicated in the etiology of other LRE, such as hepatic encephalopathy, ascites, and esophagogastric varices. Hepatic encephalopathy is a neurological complication of liver disease that is thought to be caused by the accumulation of ammonia and other toxic substances in the blood.30 ATX and LPA signaling have been shown to increase the production of ammonia in the liver and lead to enhancement of blood-brain barrier permeability,31 which can contribute to hepatic encephalopathy onset.32 Additionally, ATX and LPA signaling were found to increase blood vessel permeability in the liver, potentially leading to the leakage of fluid into the peritoneal cavity and ascites, and raised the production of angiogenic factors promoting the growth of new blood vessels towards the development of varices.33 It has also been reported that serum ATX levels are high in cirrhotic patients with ruptured varices, ascites, and encephalopathy.34 Taken together, ATX and LPA signaling are possibly involved in LRE, including HCC, hepatic encephalopathy, ascites, and esophagogastric varices, independently of liver fibrosis. Measuring serum ATX levels may therefore assist in predicting future LRE.

Lastly, previous research supports a notable gender difference in reference levels of ATX, with higher values in women.35 This discrepancy is thought to be influenced by estrogen, which reportedly regulates ATX production and secretion.36 As such, gender is an important factor to consider when interpreting ATX levels in patients. To account for these known physiological and pathological gender differences, the present cohort analysis was performed using serum ATX divided by the ULN of each gender. This approach led to a more accurate and informative assessment of prognosis as compared with analyzing the data as a whole. Indeed, considering gender is a critical point in laboratory testing of ATX and clinical decision-making.

This study had several limitations, including a retrospective nature, single-center design, and relatively small sample size. Given that the study population consisted exclusively of Japanese individuals, it will be important to validate our findings in larger cohorts of other ethnicities. Moreover, we did not evaluate the time course of serum ATX levels, which could potentially change over time. To establish the clinical significance of ATX in NAFLD, future research involving larger and more diverse patient populations that monitor the changes in serum ATX levels over time will be necessary.

In conclusion, ATX holds promise as a reliable biomarker for predicting the likelihood of adverse long-term outcomes in NAFLD, including death, HCC, and LRE. The addition of ATX as a prognostic biomarker may aid in identifying patients at higher risk of adverse outcomes, allowing for more targeted interventions and better disease management.

## Data Availability

All data produced in the present study are available upon reasonable request to the authors

## Abbreviations

AFP: alpha-fetoprotein
ATX: autotaxin
AUROC: area under the receiver operating characteristic curve
CI: confidence interval
DL: dyslipidemia
DM: diabetes mellites
HCC: hepatocellular carcinoma
HOMA-IR: homeostasis model assessment of insulin resistance
HR: hazard ratio
HT: hypertension
IQR: interquartile range
LDL-C: low density lipoprotein cholesterol
LPA: lysophosphatidic acid
LRE: liver-related events
NAFLD: non-alcoholic fatty liver disease
TC: total cholesterol
ULN: upper limit of normal

## Author contributions

Kimura T designed the research; Iwadare T, Kimura T, Okumura T, Wakabayashi S, Nakajima T, Kondo S, Kobayashi H, Yamashita Y, Sugiura A, Fujimori N, Yamazaki T, Kunimoto H, Joshita S, and Tanaka N collected clinical data; Iwadare T, Kimura T, Kondo S, Shimamoto S, and Igarashi K performed the assays; Iwadare T and Kimura T analyzed the data; Iwadare T, Kimura T, and Tanaka N wrote the paper; Umemura T supervised this study.

## Acknowledgements

The authors thank Asami Yamazaki and Mie Karakida for their assistance in sample and database preparation. We also thank Trevor Ralph for his help in English proofreading.

## References

1. Angulo P. Nonalcoholic fatty liver disease. New England Journal of Medicine 2002;346:1221–1231.

2. Diehl AM, Day C. Cause, Pathogenesis, and Treatment of Nonalcoholic Steatohepatitis. N Engl J Med 2017;377:2063–2072.

3. Tanaka N, Kimura T, Fujimori N, et al. Current status, problems, and perspectives of non-alcoholic fatty liver disease research. World journal of gastroenterology 2019;25:163.

4. Angulo P, Kleiner DE, Dam-Larsen S, et al. Liver Fibrosis, but No Other Histologic Features, Is Associated With Long-term Outcomes of Patients With Nonalcoholic Fatty Liver Disease. Gastroenterology 2015;149:389–97.e10.

5. Fujii H, Iwaki M, Hayashi H, et al. Clinical Outcomes in Biopsy-Proven Nonalcoholic Fatty Liver Disease Patients: A Multicenter Registry-based Cohort Study. Clin Gastroenterol Hepatol 2022.

6. Tsutsui M, Tanaka N, Kawakubo M, et al. Serum fragmented cytokeratin 18 levels reflect the histologic activity score of nonalcoholic fatty liver disease more accurately than serum alanine aminotransferase levels. Journal of clinical gastroenterology 2010;44:440–447.

7. Kimura T, Tanaka N, Fujimori N, et al. Serum thrombospondin 2 is a novel predictor for the severity in the patients with NAFLD. Liver International 2021;41:505–514.

8. Fujimori N, Kimura T, Tanaka N, et al. 2-Step PLT16-AST44 method: Simplified liver fibrosis detection system in patients with non-alcoholic fatty liver disease. Hepatol Res 2022;52:352–363.

9. Stracke M, Krutzsch HC, Unsworth EJ, et al. Identification, purification, and partial sequence analysis of autotaxin, a novel motility-stimulating protein. Journal of Biological Chemistry 1992;267:2524–2529.

10. Nakanaga K, Hama K, Aoki J. Autotaxin—an LPA producing enzyme with diverse functions. The journal of biochemistry 2010;148:13–24.

11. Fujimori N, Umemura T, Kimura T, et al. Serum autotaxin levels are correlated with hepatic fibrosis and ballooning in patients with non-alcoholic fatty liver disease. World Journal of Gastroenterology 2018;24:1239.

12. Joshita S, Ichikawa Y, Umemura T, et al. Serum autotaxin is a useful liver fibrosis marker in patients with chronic hepatitis B virus infection. Hepatology Research 2018;48:275–285.

13. Yamazaki T, Joshita S, Umemura T, et al. Association of serum autotaxin levels with liver fibrosis in patients with chronic hepatitis C. Scientific reports 2017;7:1–10.

14. Joshita S, Umemura T, Usami Y, et al. Serum autotaxin is a useful disease progression marker in patients with primary biliary cholangitis. Scientific reports 2018;8:1–10.

15. Komatsu M, Yazaki M, Tanaka N, et al. Citrin deficiency as a cause of chronic liver disorder mimicking non-alcoholic fatty liver disease. Journal of hepatology 2008;49:810–820.

16. Umemura S, Arima H, Arima S, et al. The Japanese Society of Hypertension guidelines for the management of hypertension (JSH 2019). Hypertension Research 2019;42:1235–1481.

17. Teramoto T, Sasaki J, Ishibashi S, et al. Diagnostic criteria for dyslipidemia executive summary of the Japan Atherosclerosis Society (JAS) guidelines for the diagnosis and prevention of atherosclerotic cardiovascular diseases in Japan—2012 version. Journal of atherosclerosis and thrombosis 2013;20:655–660.

18. Araki E, Goto A, Kondo T, et al. Japanese clinical practice guideline for diabetes 2019. Diabetology international 2020;11:165–223.

19. Gayoso-Diz P, Otero-González A, Rodriguez-Alvarez MX, et al. Insulin resistance (HOMA-IR) cut-off values and the metabolic syndrome in a general adult population: effect of gender and age: EPIRCE cross-sectional study. BMC endocrine disorders 2013;13:1–10.

20. Kleiner DE, Brunt EM, Van Natta M, et al. Design and validation of a histological scoring system for nonalcoholic fatty liver disease. Hepatology 2005;41:1313–21.

21. Bruix J, Sherman M. Management of hepatocellular carcinoma: an update. Hepatology (Baltimore, Md.) 2011;53:1020.

22. Nakagawa H, Ikeda H, Nakamura K, et al. Autotaxin as a novel serum marker of liver fibrosis. Clinica chimica acta 2011;412:1201–1206.

23. Kaffe E, Katsifa A, Xylourgidis N, et al. Hepatocyte autotaxin expression promotes liver fibrosis and cancer. Hepatology 2017;65:1369–1383.

24. Kostadinova L, Shive CL, Anthony DD. Elevated Autotaxin and LPA Levels During Chronic Viral Hepatitis and Hepatocellular Carcinoma Associate with Systemic Immune Activation. Cancers (Basel) 2019;11.

25. Honda Y, Imajo K, Kobayashi T, et al. Autotaxin is a valuable biomarker for the prediction of liver fibrosis in patients with non-alcoholic fatty liver disease. Hepatol Res 2019;49:1136–1146.

26. Sheka AC, Adeyi O, Thompson J, et al. Nonalcoholic Steatohepatitis: A Review. Jama 2020;323:1175–1183.

27. Yokomori H, Ando W, Kaneko F, et al. Autotaxin and vascular endothelial growth factor receptor-2 and -3 are related to vascular development during the progression of chronic viral hepatitis C. Apmis 2018;126:913–921.

28. Ando W, Kaneko F, Shimamoto S, et al. Long-term prediction of hepatocellular carcinoma using serum autotaxin levels after antiviral therapy for hepatitis C. Ann Hepatol 2022;27:100660.

29. Peyruchaud O, Saier L, Leblanc R. Autotaxin Implication in Cancer Metastasis and Autoimunne Disorders: Functional Implication of Binding Autotaxin to the Cell Surface. Cancers (Basel) 2019;12.

30. Komatsu M, Kimura T, Yazaki M, et al. Steatogenesis in adult-onset type II citrullinemia is associated with down-regulation of PPARalpha. Biochim Biophys Acta 2015;1852:473–81.

31. Masago K, Kihara Y, Yanagida K, et al. Lysophosphatidic acid receptor, LPA6, regulates endothelial blood-brain barrier function: Implication for hepatic encephalopathy. Biochem Biophys Res Commun 2018;501:1048–1054.

32. Roy S, Chakrabarti M, Dasgupta H, et al. Inhibition of Autotaxin Ameliorates LPA-Mediated Neuroinflammation and Alleviates Neurological Dysfunction in Acute Hepatic Encephalopathy. ACS Chem Neurosci 2022;13:2829–2841.

33. Smyth SS, Kraemer M, Yang L, et al. Roles for lysophosphatidic acid signaling in vascular development and disease. Biochim Biophys Acta Mol Cell Biol Lipids 2020;1865:158734.

34. Shao X, Uojima H, Setsu T, et al. Usefulness of autotaxin for the complications of liver cirrhosis. World J Gastroenterol 2020;26:97–108.

35. Nakamura K, Igarashi K, Ide K, et al. Validation of an autotaxin enzyme immunoassay in human serum samples and its application to hypoalbuminemia differentiation. Clin Chim Acta 2008;388:51–8.

36. Zhang G, Cheng Y, Zhang Q, et al. ATX-LPA axis facilitates estrogen-induced endometrial cancer cell proliferation via MAPK/ERK signaling pathway. Mol Med Rep 2018;17:4245–4252.

